# West Nile morbidity and mortality in a Mid-Atlantic healthcare system, 2013-2024

**DOI:** 10.1101/2025.06.18.25329873

**Authors:** Seth D. Judson, Paul Auwaerter, David Dowdy

**Author notes:** Corresponding author: Seth D. Judson, 1830 E Monument St, Rm 450B, Baltimore, MD 21205.

## Abstract

We investigated West Nile cases in a Mid-Atlantic healthcare system, finding significant morbidity and mortality from neuroinvasive disease, notably among immunocompromised patients. Encephalitis was the most common complication and was associated with increased age, alcohol use disorder, immunocompromised status, and a higher Charlson Comorbidity Index.

## Introduction

West Nile virus (WNV) causes the greatest morbidity and mortality of any vector-borne virus in North America [1]. There are currently no treatments or vaccines for WNV in humans; therefore, preventing infection is the primary method for mitigating disease. Understanding the local disease burden and risk factors for severe WNV disease can help clinicians and public health practitioners develop an appropriate response.

Approximately 25% of patients infected with WNV develop a febrile syndrome called West Nile fever (WNF), while <1% develop neuroinvasive disease (WNND) [2,3]. The main manifestations of WNND include meningitis (WNM), encephalitis (WNE), and acute flaccid paralysis (AFP) [4]. These complications may coincide and can result in lifelong neurologic sequelae and death, with a case fatality rate of ∼10% [1].

While there have been multiple studies of the burden and risk factors for WNND in North America, most of these studies are over a decade old and primarily included patients from the western and midwestern United States [5,6]. Considering the heterogeneity among patient populations, increasing prevalence of immunosuppressive treatments, and novel data available in electronic medical records (EMR), contemporary analyses of WNND and mortality are needed. We aimed to characterize the burden of, and risk factors for, severe WNND and WNV mortality from 2013 to 2024 within an academic healthcare system in the Mid-Atlantic region.

## Methods

### Ethical Considerations

This retrospective cohort study of Johns Hopkins Healthcare System (JHHS) patients was approved by the Johns Hopkins School of Medicine Institutional Review Board (IRB00488802) under a waiver of informed consent. This study was a secondary analysis of existing data that did not involve intervention or interaction with human subjects.

### Cohort selection

The JHHS includes more than 40 outpatient facilities and 6 hospitals in Maryland, Washington, DC, and Florida (which has only pediatric facilities and was excluded from this analysis). Data were extracted from the Epic Clarity database that includes both inpatient and outpatient EMRs using search criteria for patients with ICD-10 codes for WNV infection (A92.30, A92.31, A92.32, A92.39) or positive WNV diagnostic results (WNV IgG, IgM, or PCR). Data from JHHS were accessed on 4/24/2025. The final cohort included cases that occurred between January 1, 2013 (the year that Epic was launched at JHHS) and December 31, 2024. All records identified with positive ICD-10 codes and/or laboratory results for WNV underwent chart review for inclusion using the CDC’s West Nile virus neuroinvasive and non-neuroinvasive probable/confirmed case definitions to identify cases of WNND and WNF [7].

Inclusion criteria included adults (≥18 years of age) diagnosed with laboratory-established WNV infection (positive serum and/or CSF WNV IgM or PCR) in the setting of compatible clinical history between 2013 and 2024. Clinical signs/symptoms, laboratory results, and imaging were used to classify patients as having WNF and WNND, defined as WNE, WNM, and/or AFP (S Table 1).

### Outcomes and Covariates

Our primary outcome for morbidity was WNND. Given that WNE is the most prevalent severe form of WNND, we assessed risk factors for WNE in a secondary analysis.

Our co-primary outcomes for mortality were all-cause mortality and mortality directly attributable to WNV. Our secondary outcomes included hospitalization, ICU admission, and hospital length of stay (LOS).

We assessed potential risk factors with a priori evidence of increased risk for WNND and mortality [5,6,8]. We also calculated the Charlson Comorbidity Index (CCI) for each patient as a composite risk factor (excluding age from the calculation). The CCI was calculated using the ICD-10-based scoring system [9]. Comorbidities in the CCI and risk factors were assessed at the date of the encounter for WNV. We also evaluated immunocompromise as a risk factor, including history of solid organ transplant, active malignancy, HIV with CD4 count <200, on immunosuppressants (calcineurin inhibitors, high-dose prednisone, mycophenolate, and/or biologics), or receiving chemotherapy.

### Statistical Analysis

We calculated descriptive statistics for our primary and secondary outcomes. For our risk factor analysis for WNE, we first used logistic regression to assess bivariate associations with each outcome. Covariates with a p-value <0.2 and present in >5 patients total were entered into a multivariable logistic regression model, and stepwise backward selection was then used to identify the model with the optimal AIC (Akaike Information Criterion) value using the MASS package in R. Given the potential for collinearity between covariates and the CCI, a separate multivariable model containing age, sex, and CCI was created. Goodness-of-fit for final models was assessed using the Hosmer-Lemeshow test with five bins, and multicollinearity was assessed using Variance Inflation Factors (VIFs). A two-sided p-value <0.05 was used for statistical significance for the final covariates. All statistical analyses were conducted in R version 4.4.2.

## Results

Among 367 adult patients with positive ICD-10 codes and/or lab results for WNV, 102 met CDC confirmed/probable case definitions. Our final cohort included 86 patients with WNV from 2013-2024. Most cases (78/86, 91%) were diagnosed with WNV during August through October.

### West Nile Neuroinvasive Disease

Among the 86 patients in the cohort, 71 (83%) had WNND and 15 (17%) had WNF (Table 1). Of those with WNND, 47 (66%) had encephalitis/meningoencephalitis (i.e. WNE), 18 (25%) had meningitis without encephalitis, 4 (6%) had AFP, and 2 (3%) had transverse myelitis. The majority of patients with WNND were male (47/71, 66%), ≥ 50 years old (57/71, 80%), and had a CCI >0 (47/71, 66%). Of 21 immunocompromised patients, 20 (95%) developed WNND, including 16/17 (94%) on immunosuppressants (including calcineurin inhibitors, mycophenolate, prednisone, biologics [adalimumab, rituximab, ocrelizumab], interferon beta 1a, and antineoplastics [venetoclax and acalabrutinib]), and 7/7 (100%) with history of solid organ transplant.

**Table 1.**
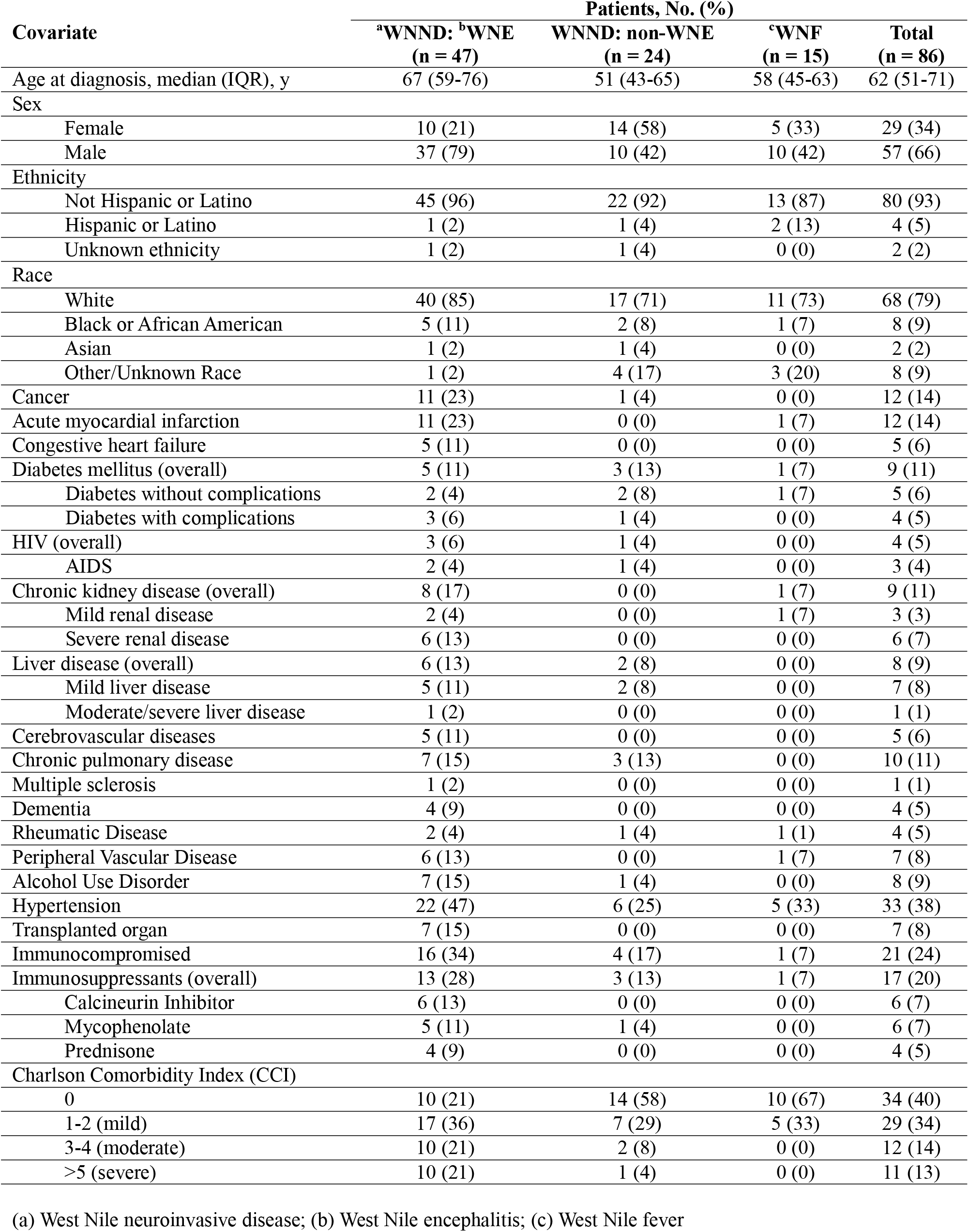
Characteristics of patients with WNV in a Mid-Atlantic healthcare system, 2013-2024.

Potential risk factors for WNE (p<0.2) in the univariable analysis included age, male sex, cancer, acute myocardial infarction, chronic kidney disease (CKD), peripheral vascular disease, hypertension, alcohol use disorder, immunosuppressant use, immunocompromised status, and CCI. The most parsimonious model (lowest AIC) included age (adjusted odds ratio, aOR, 2.46 per 10 years, 95% confidence interval 1.64-4.05), CKD (aOR 6.76, 0.76-175), alcohol use disorder (aOR 11.8, 1.59-248), and immunocompromised status (aOR 6.71, 1.55-38.2) (S Table 2). In the multivariable model including CCI, indices of 3-4 (aOR 7.84, 1.28-75.1) and >5 (aOR 19.9, 2.78-421) relative to an index of 0 were significantly associated with WNE (S Table 3). In both multivariable models, variance inflation factors were all below 2, indicating no significant collinearity, and the Hosmer-Lemeshow test demonstrated adequate fit (p-values 0.61 and 0.29).

### Hospitalization

Of the 86 patients with WNV in the cohort, 74 (86%) were hospitalized, including 68/71 (96%) of patients with WNND and 6/16 (40%) with WNF. Of the 74 admitted patients, 26 (35%) required ICU admission (all with WNND), and 21 (28%) were immunocompromised. The overall median hospital LOS was 8 days (interquartile range 5-15).

### Mortality

Considering all-cause mortality, 9/86 (10%) of patients with WNV infection from 2013-2024 died; all of these patients had WNE, and two also had AFP. Among these patients, 5 (55%) died within 30 days of admission, with WNV identified as the cause of death. The CFR directly attributable to WNND was 5/71 (7%). All of those who died were > 50 years old and had a CCI > 0; 4/9 (44%) of those who died were immunocompromised (S Table 4).

## Discussion

We identified significant morbidity and mortality from WNV in a Mid-Atlantic healthcare system over the past decade. The majority of diagnosed patients had WNND, with WNE being the most common neurologic manifestation, aligning with prior studies [3]. The high proportions of hospitalization (86%), ICU admission (35%), and prolonged LOS (median 8 days) further support recent findings of the impact of WNV on healthcare resources [10]. Of nine patients who died, WNV was identified as the cause of death in five–all of whom died within 30 days of diagnosis.

Our study also reinforces the known association between increased age and WNND, with the odds of WNE doubling with every decade of age in our cohort. Another notable finding in our study was the high prevalence of WNND among immunocompromised individuals who were diagnosed with WNV. Immunocompromise was a strong risk factor for WNE, including the use of various immunosuppressants, which corroborates recent findings that immunosuppression is associated with the severity of WNND [11]. Furthermore, alcohol use disorder was identified as a significant risk factor for WNE, consistent with prior studies [6,12].

A limitation of this analysis is the inclusion of probable WNV cases, which were diagnosed serologically and could represent cross-reactivity with other flaviviruses. Using CDC case definitions for acute (probable/confirmed) WNV infection also limited our sample size and statistical power for certain covariates. In this setting, we found that the CCI may be a useful tool for identifying the risk of WNE, as it can be readily calculated from clinical data when individual comorbidities are less prevalent.

Overall, our findings of substantial morbidity and mortality for WNV in a Mid-Atlantic healthcare system over the past decade highlight the need for increased surveillance, targeted prevention, and countermeasure development. These measures, in particular a WNV vaccine, could be especially impactful among the immunocompromised population and older patients with multiple comorbidities.

## Data Availability

Data not publicly available

## Acknowledgments

This work was supported by the National Institute of Health [T32 AI007291-34] to SDJ and the Fisher Center for Environmental Infectious Diseases. Author contributions: Writing—original draft: S.D.J. Writing—review and editing: S.D.J., D.D., P.G.A. The authors deny any conflicts of interest. Data not publicly available.

