## Supplemental Appendix for "West Nile morbidity and mortality in a Mid-Atlantic healthcare system, 2013-2024"

| S Table 1. Diagnostic Criteria for West Nile Neuroinvasive Disease Outcomes | |
| --- | --- |
| Diagnosis | Criteria |
| WNF | Positive serum IgM or PCR  AND  Any of the following clinical signs/symptoms:  Fever, myalgias, fatigue, rash, nausea/vomiting/diarrhea, headache  Cannot have: neurological signs/symptoms, or positive CSF IgM or PCR |
| WNM | Positive serum/CSF IgM or PCR  AND  Signs/symptoms of meningeal inflammation (nuchal rigidity, neck pain, photophobia, headache), CSF pleocytosis, and/or consistent MRI imaging |
| WNE | Positive serum/CSF IgM or PCR  AND  Altered mental status, seizures, abnormal EEG, focal neurological deficits, and/or consistent MRI imaging |
| AFP | Positive serum/CSF IgM or PCR  AND  Acute onset of flaccid limb weakness or paralysis, consistent MRI imaging/EMG and/or examination and diagnosis of AFP by neurologist |

| **S Table 2. Univariable and Multivariable Associations with WNE** | | | |
| --- | --- | --- | --- |
| **Covariate** | **Unadjusted OR (95% CI)** | **Adjusted OR (95% CI)*** | **P-value^†^** |
| Age per decade | 1.93 (1.40 to 2.83) | 2.46 (1.64 to 4.05) | <0.0001 |
| Male sex | 3.51 (1.40 to 9.28) | - | - |
| Cancer | 11.6 (2.10 to 218) | - | - |
| Acute myocardial infarction | 11.6 (2.10 to 218) | - | - |
| Chronic kidney disease | 7.79 (1.34 to 148) | 6.76 (0.756 to 175) | 0.14 |
| Peripheral vascular disease | 5.56 (0.89 to 108) | - | - |
| Hypertension | 2.24 (0.92 to 5.67) | - | - |
| Alcohol use disorder | 6.65 (1.11 to 127) | 11.84 (1.59 to 248) | 0.035 |
| Immunosuppressants | 3.35 (1.06 to 12.8) | - | - |
| Immunocompromised | 3.51 (1.21 to 11.8) | 6.71 (1.55 to 38.2) | 0.018 |
| **OR= Odds Ratio; CI= Confidence Interval; Adjusted results based on multivariable logistic regression  †P-value for adjusted results | | | |

AIC of the initial model including all of the above covariates: 97.5

AIC of the final model using backwards selection (including age, CKD, alcohol use disorder, and immunocompromised): 90.8

| **S Table 3. Univariable and Multivariable Associations of CCI with WNE** | | | |
| --- | --- | --- | --- |
| **Covariate** | **Unadjusted OR (95% CI)** | **Adjusted OR (95% CI)*** | **P-value^†^** |
| Age per decade | 1.93 (1.40 to 2.83) | 1.97 (1.35 to 3.16) | <0.0001 |
| Male sex | 3.51 (1.40 to 9.28) | 2.07 (0.615 to 7.13) | 0.24 |
| CCI  0  1-2  3-4  >5 | Ref  3.40 (1.22 to 9.98)  12.0 (2.60 to 88.0)  24.0 (3.86 to 471) | Ref  2.06 (0.608 to 7.06)  7.84 (1.28 to 75.1)  19.9 (2.78 to 421) | Ref  0.24  0.04  0.01 |
| **OR= Odds Ratio; CI= Confidence Interval; Adjusted results based on multivariable logistic regression  †P-value for adjusted results | | | |

| **S Table 4. Characteristics of patients with WNV and mortality, 2013-2024** | | | |
| --- | --- | --- | --- |
| **Covariate** | **Patients, No. (%)** | | |
|  | **WNV & no death**  **(n =77)** | **WNV & death**  **(n = 9)** | **Total**  **(n = 86)** |
| Age at diagnosis, median (IQR), y | 61 (50-71) | 68 (55-74) | 62 (51-71) |
| Sex |  |  |  |
| Female | 28 (36) | 1 (11) | 29 (34) |
| Male | 49 (64) | 8 (89) | 57 (66) |
| Ethnicity |  |  |  |
| Not Hispanic or Latino | 71 (92) | 9 (100) | 80 (93) |
| Hispanic or Latino | 4 (5) | 0 (0) | 4 (5) |
| Unknown ethnicity | 2 (3) | 0 (0) | 2 (2) |
| Race |  |  |  |
| White | 61 (80) | 7 (78) | 68 (79) |
| Black or African American | 7 (9) | 1 (11) | 8 (9) |
| Asian | 2 (3) | 0 (0) | 2 (2) |
| Other/Unknown Race | 7 (9) | 1 (11) | 8 (9) |
| Cancer | 12 (15) | 0 (0) | 12 (14) |
| Acute myocardial infarction | 7 (9) | 5 (55) | 12 (14) |
| Congestive heart failure | 3 (4) | 2 (22) | 5 (6) |
| Diabetes mellitus (overall) | 9 (11) | 0 (0) | 9 (11) |
| Diabetes without complications | 5 (7) | 0 (0) | 5 (6) |
| Diabetes with complications | 4 (5) | 0 (0) | 4 (5) |
| HIV (overall) | 4 (5) | 0 (0) | 4 (5) |
| AIDS | 3 (4) | 0 (0) | 3 (4) |
| Chronic kidney disease (overall) | 4 (5) | 5 (56) | 9 (11) |
| Mild renal disease | 2 (3) | 1 (11) | 3 (3) |
| Severe renal disease | 2 (3) | 4 (44) | 6 (7) |
| Liver disease (overall) | 5 (7) | 3 (33) | 8 (9) |
| Mild liver disease | 5 (7) | 2 (22) | 7 (8) |
| Moderate/severe liver disease | 0 (0) | 1 (11) | 1 (1) |
| Cerebrovascular diseases | 2 (3) | 3 (33) | 5 (6) |
| Chronic pulmonary disease | 8 (10) | 2 (22) | 10 (11) |
| Multiple sclerosis | 1 (1) | 0 (0) | 1 (1) |
| Dementia | 4 (5) | 0 (0) | 4 (5) |
| Rheumatic Disease | 4 (5) | 1 (4) | 4 (5) |
| Peripheral Vascular Disease | 5 (7) | 2 (22) | 7 (8) |
| Alcohol Use Disorder | 6 (8) | 2 (22) | 8 (9) |
| Hypertension | 28 (36) | 5 (57) | 33 (38) |
| Transplanted organ | 3 (4) | 4 (44) | 7 (8) |
| Immunocompromised | 17 (22) | 4 (44) | 21 (24) |
| Immunosuppressants (overall) | 13 (17) | 4 (44) | 17 (20) |
| Calcineurin Inhibitor | 3 (4) | 3 (33) | 6 (7) |
| Mycophenolate | 2 (3) | 4 (4) | 6 (7) |
| Prednisone | 1 (1) | 3 (33) | 4 (5) |
| Charlson Comorbidity Index (CCI) |  |  |  |
| 0 | 34 (44) | 0 (0) | 34 (40) |
| 1-2 (mild) | 27 (35) | 2 (22) | 29 (34) |
| 3-4 (moderate) | 8 (10) | 4 (44) | 12 (14) |
| >5 (severe) | 8 (10) | 3 (33) | 11 (13) |
